# Body Size and Composition Trends in U.S. Adults with a History of Cancer: NHANES 1999–2018

**DOI:** 10.1101/2025.07.26.25332243

**Authors:** Benjamin Brockbank, Rand Talal Akasheh, Yevgeniya Gokun, Faiza Kalam, Cuthbert Mario Mahenge, Electra D. Paskett, Ting-Yuan David Cheng

**Affiliations:** Brigham Young University, Simmons Center for Cancer Research, Provo, UT, United States; Division of Cancer Prevention and Control, Department of Internal Medicine, College of Medicine, The Ohio State University, Columbus, OH, United States; Ohio State University Comprehensive Cancer Center, The Ohio State University, Columbus, OH, United States; Center for Biostatistics, The Ohio State University College of Medicine, Columbus, OH, United States

**Author notes:** **Corresponding author:** Ting-Yuan David Cheng, PhD, Division of Cancer Prevention and Control, Department of Internal Medicine, College of Medicine, The Ohio State University, 3650 Olentangy River Rd, Suite 200, Columbus, OH 43214, United States.

**Keywords:** Body composition, dual-energy X-ray absorptiometry, National Health and Nutrition Examination Survey, cancer survivors

## Abstract

**Purpose:** High fat mass and low muscle mass are associated with increased mortality and risk of second primary cancers in cancer survivors. This longitudinal study examined trends in body size and composition among US adults with a history of cancer from 1999 to 2018.

**Methods:** Data from self-reported cancer survivors aged ≥20 years in the Continuous National Health and Nutrition Examination Survey 1999-2006 and 2011-2018 were analyzed. Outcome measurements included body mass index (BMI, n=3,544) waist circumference (WC, n=3,354), and DEXA-derived indices, including fat mass index (FMI, n=1,767), total lean mass (LM, n=1,775), and appendicular skeletal muscle mass index (ASMMI, n=1,741). Multivariate linear regressions were used to analyze across data cycle associations, adjusting for age, sex, and race/ethnicity.

**Results:** Across 19 years, average BMI increased from 27.1 kg/m² to 30.0 kg/m² (p<.001) and average WC increased from 96.1 cm to 103.8 cm (p<.001) among all cancer survivors and increased in Hispanic and non-Hispanic White subgroups, but not non-Hispanic Black or other race/ethnicity subgroups. Total LM increased overall between 1999 and 2006, from 46.6 kg to 49.5 kg (p=.002), and increased in the non-Hispanic White subgroup (p=.009). FMI and ASMMI showed no significant overall changes, although FMI levels remained elevated.

**Conclusion:** US cancer survivors experienced notable increases in BMI and WC from 1999 to 2018, with LM gains observed in some racial/ethnic groups, particularly from 1999 to 2006.

**Implications for Cancer Survivors:** These findings highlight the need for targeted interventions addressing obesity and fat mass to improve long-term health outcomes among cancer survivors.

## Introduction

During the past decade, cancer survivors and patients in the United States (US) have exhibited a higher obesity prevalence (body mass index [BMI] ≥30 kg/m^2^) compared with the general US population.^1–4^ The high obesity prevalence in cancer survivors aligns with emerging evidence linking obesity to an increased incidence of metabolically and hormonally driven cancers, such as breast and colorectal cancer.^5,6^ High BMI is associated with a significantly elevated risk of developing secondary cancers^7^ and mortality among cancer survivors.^8–10^ While studies providing this evidence have evaluated BMI as the primary measure, BMI has significant limitations as it does not distinguish between fat and lean mass.^11^ This limitation has led to the observation of an “obesity paradox” in which lower cancer mortality rates are identified in the overweight to low-obese range,^12,13^ potentially due to a greater relative contribution of muscle mass than fat mass in these individuals.^14^ High fat mass and low muscle mass are associated with higher all-cause mortality in cancer survivors,^14–16^ highlighting the need for comprehensive body composition assessment.

To date, very little research has evaluated the longitudinal pattern of body composition measures in cancer survivors. To address this gap, the current study analyzed longitudinal data (1999-2006 and 2011-2018) from dual-energy X-ray absorptiometry (DEXA), a method that employs low-energy X-rays to estimate the lean mass and fat mass of the total body,^11^ and anthropometric data in a nationally representative sample of cancer survivors. Our goal was for the study findings to inform targeted prevention and treatment strategies to improve the health and nutritional status of cancer survivors.

## Methods

### Study Participants

The National Health and Nutrition Examination Survey (NHANES) is a nationally representative cross-sectional survey conducted by the Centers for Disease Control and Prevention National Center for Health Statistics, which assesses the health of non-institutionalized individuals in the US.^17^ NHANES uses a complex, multistage sampling design to recruit approximately 10,000 non-institutionalized individuals every 2 years. Data from continuous NHANES 2-year cycles from 1999 to 2006 and then from 2011 to 2018 were included in this study. We excluded two 2-year cycles (2007-2008 and 2009-2010) in which whole-body DEXA was not performed. The 2017-2018 cycle represents the most recently released and complete NHANES dataset available for all measures. Trained staff conducted interviews and used standardized methods to obtain physical measurements under identical conditions at a mobile examination center, which is brought to selected communities. The survey response rate decreased from 85% in 2000 to 48% in 2018.^18^

This study was deemed to be non-human subject research by the Institutional Review Board at the Ohio State University as the data were de-identified.

### Outcomes of Interest

All participants aged 2 years or older and able to stand underwent NHANES anthropometry procedures.^19^ Height in meters was assessed while standing, using a stadiometer with an adjustable headpiece and a fixed backboard. Weight in kilograms was calculated using a digital weight scale, with the participant wearing only a gown and slippers. BMI was then computed as weight (kg) divided by height squared (m^2^). Waist circumference (WC) was measured in centimeters with a measuring tape placed just above the iliac crest.^19^

Certified radiology technologists used Hologic QDR 4500A fan-beam bone densitometers to provide whole-body DEXA measurements.^20^ All pregnant individuals, determined through a questionnaire and confirmed by a positive pregnancy test, were excluded from DEXA evaluation. Fat mass index (FMI) was calculated as the DEXA value of total body fat excluding bone mineral content divided by height squared (kg/m^2^). Total lean mass (LM) in kg was calculated as LM excluding bone mineral content. Finally, appendicular skeletal muscle mass index (ASMMI) was calculated as the sum of lean mass excluding bone mineral content of the arms and legs, divided by height squared (kg/m^2^). From 1999 to 2006, NHANES performed DEXA examinations for all participants aged 8 years or older. However, beginning in 2011, the DEXA examination was only completed for individuals aged 8 through 59 years, excluding those aged 60 or older. The 1999-2006 DEXA data underwent sequential regression multivariable imputation to address non-random missing values, allowing for confident statistical analysis.^21^ Cancer history was determined with the question, “Have you ever been told by a doctor or other health professional that you had cancer or a malignancy of any kind?” Respondents who answered “yes” were included as cancer survivors in this study.

### Covariates

Demographic data regarding age (in years at the time of examination), sex (male or female), and race and ethnicity were self-reported, collected via standardized questionnaires during in-person interviews. Race and ethnicity were categorized as Hispanic (Mexican American or other Hispanic), non-Hispanic Black or African American, non-Hispanic White, or other. Age was dichotomized as 20–59 years or ≥60 years, primarily because the DEXA variables were unavailable for ages ≥60 in the 2011–2018 NHANES cycles. Smoking status was based on two questions: whether the respondent smoked at least 100 cigarettes in their entire life (yes or no); and whether the respondent now smoked cigarettes (every day, some days, or not at all). Current smokers were defined as participants who indicated that they smoked at least 100 cigarettes in their entire life and now smoke some days or every day. Former smokers were defined as those who indicated that they smoked at least 100 cigarettes but did not currently smoke. Never smokers were those who indicated that they had not smoked at least 100 cigarettes in their entire lives. Physical activity was evaluated in NHANES using the Global Physical Activity Questionnaire.^22,23^ Components of the Physical Activity Questionnaire and the Physical Activity Questionnaire Individual Activities File were used to estimate the amount of physical activity for each participant in metabolic equivalent (MET) minutes per week, calculated by multiplying the total weekly duration of recreational, occupational, and transportation physical activities by a MET value of 4 or 8, assigned for moderate or vigorous intensity activities, respectively. The following equation was used:

*MET-min/week = [4 × weekly moderate physical activity min] + [8 × weekly vigorous physical activity min]*

The Physical Activity Guidelines for Americans^24^ were used to stratify participants according to their physical activity levels into inactive (<240 MET-min/week), not meeting guidelines (240 to <600 MET-min/week), meeting guidelines (600–1200 MET-min/week), and exceeding guidelines (>1200 MET-min/week).

### Statistical Analysis

All analyses were performed with SAS, version 9.4 (SAS Institute Inc.; Cary, NC), using population-based sampling weights (WTMEC2YR) to account for the complex survey design of NHANES. Descriptive statistics included weighted percentages and 95% confidence intervals (CIs) for categorical variables.

For each DEXA outcome, separate models were performed for the 1999–2006 (imputed data) and 2011–2018 (non-imputed data) cycles. For the 1999–2006 data, effect estimates and standard errors (SEs) were initially obtained separately for each imputed dataset. The effect estimates included means that were averaged across imputations. SEs were pooled across imputations using within-imputation and between-imputation variances as proposed by Rubin and Schenker.^25,26^

Multivariable linear regressions were used to analyze each of the body composition measurements across 2-year data cycles, adjusting for age (continuous), sex, and race and ethnicity. In addition to the models for the overall cohort, stratification models were included for binary age, sex, and race and ethnicity, given known differences in body composition across these demographic factors.^2^ The stratification allowed for a more precise understanding of subgroup-specific trends and potential disparities over time.

Sensitivity analyses were conducted by additionally adjusting smoking status and physical activity levels in the regression models. Because not all participants received a DEXA examination, we examined the trends of BMI and WC among individuals who had DEXA data, that is, non-missing FMI. Additionally, we examined the trend of height in the BMI calculation. Several cancer-related variables were derived. We explored obesity-associated cancer and non–obesity-associated cancer as characteristics of the participants. The classification was based on research by the Centers for Disease Control and Prevention.^27^ The median time from the first cancer diagnosis to the interview was estimated using the variables age at the first cancer diagnosis and age at the interview. Years since cancer diagnosis, as well as cancer type, were included as covariates, along with age, NHANES data cycle, sex, and race and ethnicity, with BMI as the outcome variable. All tests were two-tailed, with p<0.05 indicating statistical significance.

## Results

### Participant Characteristics

Participants included in this analysis met the criteria of being non-pregnant and 20 years of age or older with a history of cancer and having BMI, WC, or at least one of the DEXA body composition indices (**Supplemental Figure 1**). **Supplemental Table 1** gives the characteristics of participants with available body size measurements (BMI and WC). Overall, 57.3% were female; 4.8% self-identified as Hispanic, 5.4% as non-Hispanic Black, 86.2% as non-Hispanic White, and 3.6% as other race or ethnicity. Additionally, 62.5% of participants were 60 years or older, and 37.5% were 20–59 years of age.

**Supplemental Table 2** compares participants with available BMI, WC, and DEXA data. Due to NHANES’s exclusion of participants aged 60 years or older from DEXA assessment in cycles 2011 to 2018, the age distribution differed between samples: only 50.7% of participants with DEXA data were older than 60 years compared with 73.5% of the participants with BMI data. Other characteristics were similar among these three samples. Breast cancer in female survivors and prostate cancer in male survivors were the most prevalent cancers, followed by non-melanoma skin cancer (**Supplemental Table 3**).

### BMI

For all cancer survivors, the adjusted mean BMI increased significantly during the 19-year period, from 27.1 kg/m^2^ (95% CI: 25.9–28.3) in 1999-2000 to 30.0 kg/m^2^ (95% CI: 29.4–30.6) in 2017-2018, with an average increase of 0.23 kg/m^2^ per 2-year data cycle (P<.001). In addition, BMI increased significantly when stratified by age group: an average increase of 0.20 kg/m^2^ in the 20–59 years cohort (P=.002), and an average increase of 0.23 kg/m^2^ in the ≥60 years cohort (P<.001) (**Figure 1**). Significant BMI increases were also observed for both male (P<.001) and female (P<.001) survivors. BMI increased significantly for Hispanic and non-Hispanic White survivors, but not for non-Hispanic Black survivors or other race and ethnicity groups (**Supplemental Table 4**).

**Figure 1.**
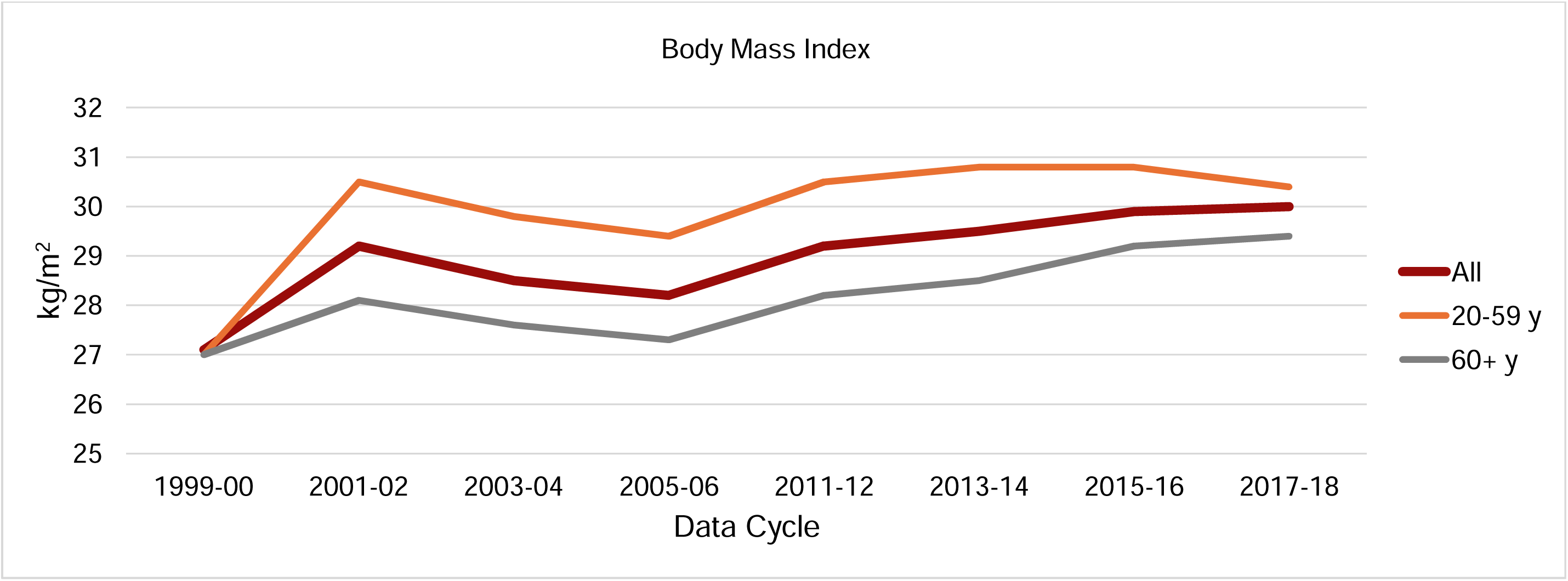
Adjusted^a^ body mass index averages: estimates overall and by age group^b^ in NHANES 1999 to 2018. ^a^ Body mass index estimates were adjusted for age (continuous), race and ethnicity, and sex. ^b^ All (includes all cancer survivors in the analysis, aged ≥20 years): P < .001 and β = .233 per cycle. Age group 20–59 years: P = .002 and β = .204. Age group 60+ years: P < .001 and β = .232.

### WC

WC changes were similar to those observed for BMI. Overall, WC increased significantly from 1999–2000 to 2017–2018, starting at 96.1 cm (95% CI, 93.2–99.1) and increasing to 103.8 cm (95% CI, 102.2–105.5), with an average increase of 0.60 cm per cycle (P<.001). These significant increases were observed in both age and sex groups (all P<.001; **Supplemental Table 5** and **Figure 2**). The Hispanic and non-Hispanic White groups exhibited significant increases in WC (both P<.001). Non-Hispanic Black survivors and other race groups had no significant change over the 19-year period.

**Figure 2.**
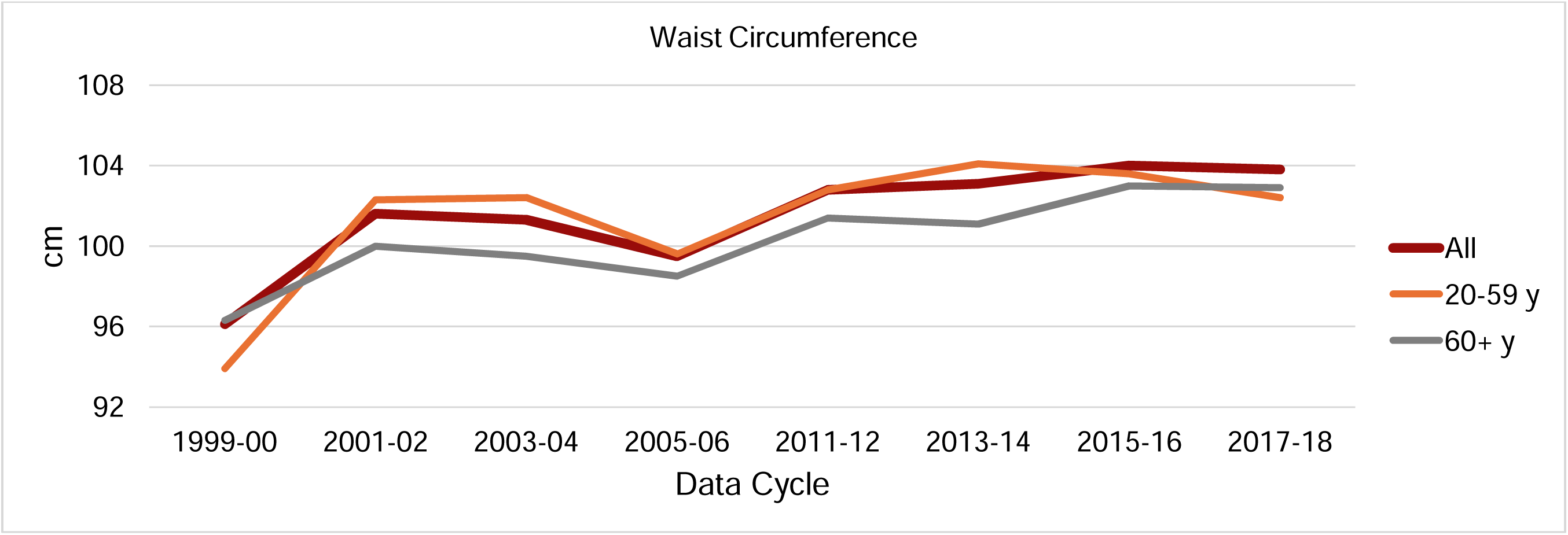
Adjusted^a^ waist circumference averages: estimates overall and by age group^b^ in NHANES 1999 to 2018. ^a^ Estimates were adjusted for age (continuous), race and ethnicity, and sex. ^b^ All (includes all cancer survivors in the analysis, aged ≥20 years): P < .001 and β = .599 per cycle. Age group 20–59 years: P <.001 and β = .523. Age group 60+ years: P < .001 and β = .582.

### FMI

Overall, FMI did not significantly change from the 1999–2000 to 2005–2006 data cycles, from 10.1 kg/m^2^ (95% CI: 9.32–10.9) to 10.9 kg/m^2^ (95% CI: 10.1–11.6), with an average increase of 0.17 kg/m^2^ (P=.23) (**Figure 3**). Additionally, FMI did not change significantly for either sex (P=.92 for male, P=.18 for female) (**Supplemental Table 6)**. However, FMI decreased for Hispanic ethnicity from 10.1 kg/m^2^ (95% CI: 9.44–10.82) in 1999–2000 to 9.56 kg/m^2^ (95% CI: 9.08–10.0) in 2005–2006, with an average decrease of 0.23 kg/m^2^ per cycle (P=.049). FMI showed no significant change among non-Hispanic Black survivors, with an average increase of 0.53 kg/m^2^ per cycle (P=.053), nor among non-Hispanic White survivors (P=.17). Similarly, FMI did not significantly change from 1999–2000 to 2005–2006 for either age cohort (P =.38 for 20–59 years of age and P=.78 for ≥60 years of age) (**Figure 3**).

**Figure 3.**
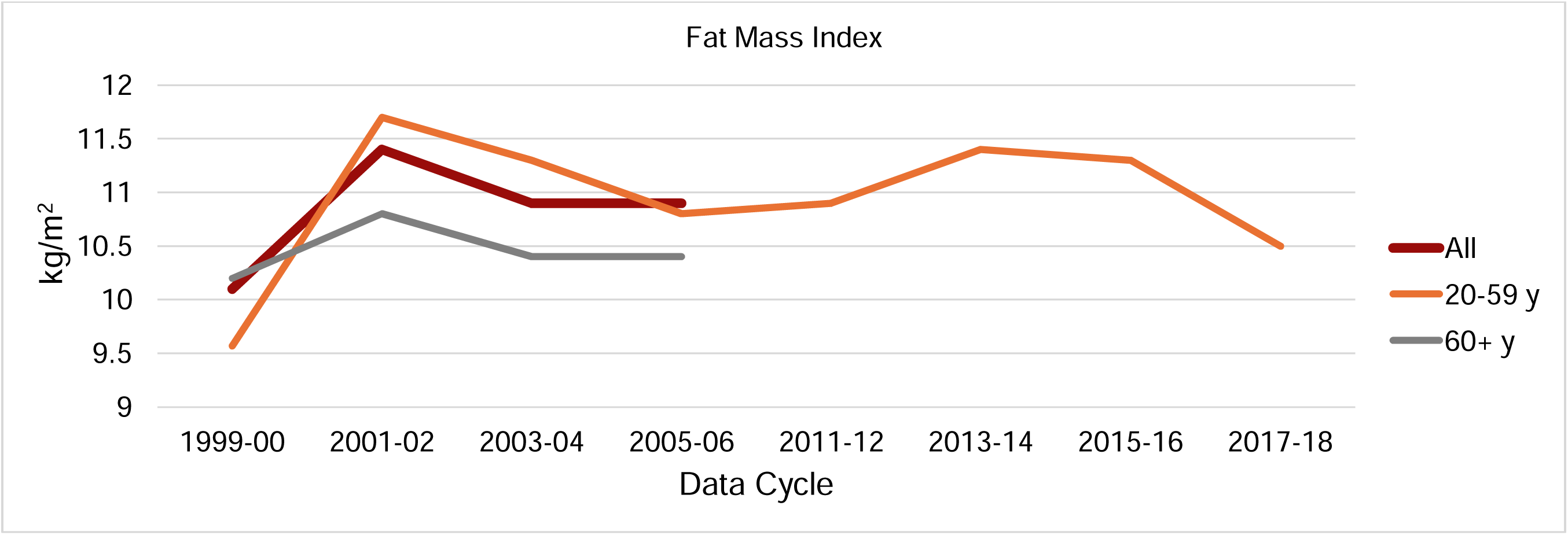
Adjusted^a^ fat mass index averages: estimates overall and by age group^b^ in NHANES 1999 to 2018. ^a^ Estimates were adjusted for age (continuous), race and ethnicity, and sex. ^b^ All (includes all cancer survivors in the analysis, aged ≥20 years 1999–2006): P = .23 and β = .172 per cycle. Age group 20–59 years, 1999–2006: P = .38 and β = .191; 2011–2018: P = .69 and β = −.121. Age group 60+ years, 1999–2006: P = .78 and β = .044.

In the later data cycles, there were no significant changes in FMI for the overall cohort or when stratified by sex or race and ethnicity from 2011–2012 to 2017–2018 (**Supplemental Table 7**).

### Total LM

LM for the overall age group increased significantly, from 46.6 kg (95% CI: 45.0– 48.2) in 1999–2000 to 49.5 kg (95% CI: 47.9–51.1) in 2005–2006, with a mean increase of 0.82 kg per cycle (P=.002) (**Figure 4**). Both sexes displayed significant increases in LM, with an average increase of 0.96 kg per cycle for male survivors (P=.04), and of 0.66 kg per cycle (P=.03) for female survivors (**Supplemental Table 8**). Among non-Hispanic White survivors, there was a significant increase from 46.8 kg (95% CI: 45.8–47.8) in 1999–2000 to 49.7 kg (95% CI, 48.0–51.3) in 2005–2006, with an average increase of 0.74 kg per cycle (P=.009). Neither Hispanic nor non-Hispanic Black survivors showed significant changes in LM from 1999–2000 to 2005–2006 (P=.06 and P=.09, respectively). There was also no significant change for either age group.

**Figure 4.**
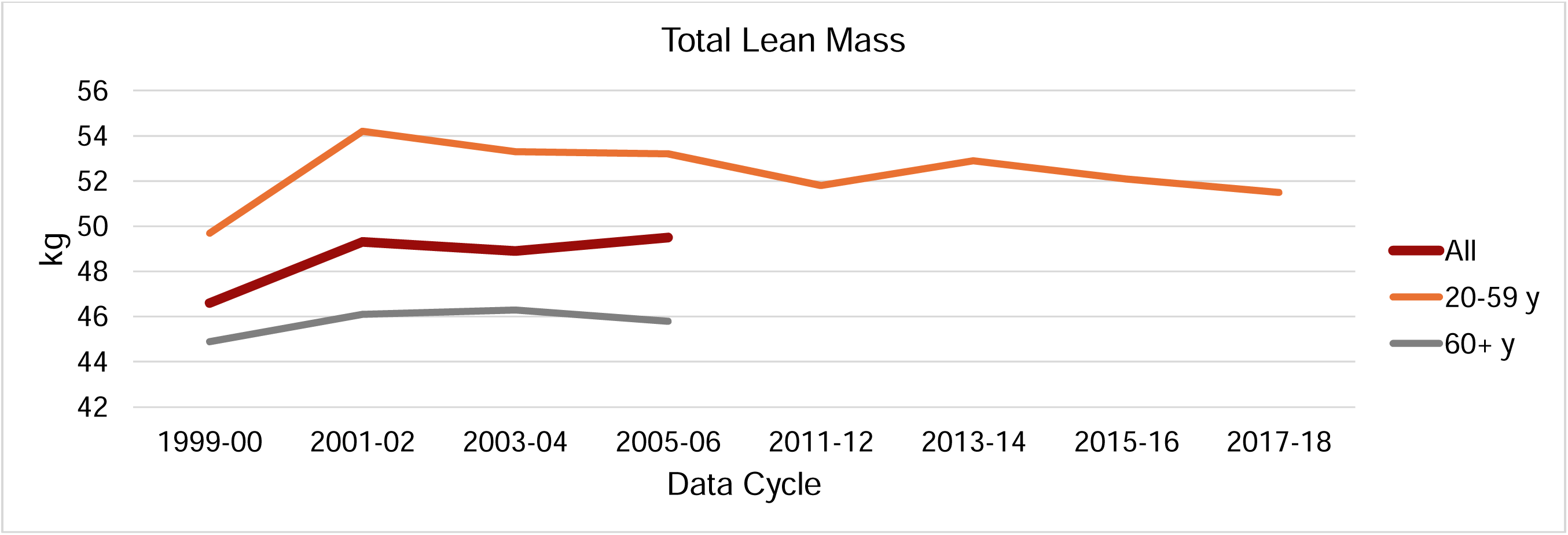
Adjusted^a^ lean mass averages: estimates overall and by age group^b^ in NHANES 1999 to 2018. ^a^ Estimates were adjusted for age (continuous), race and ethnicity, and sex. ^b^ All age group, (includes all cancer survivors in the analysis, aged ≥20 years, 1999–2006): P = .002 and β = .824 per cycle. Age group 20–59 years, 1999–2006: P = .09 and β = .730; 2011–2018: P = .814 and β = −.150. Age group 60+ years, 1999–2006: P = .26 and β = .43.

For later cycles from 2011–2012 to 2017–2018 for the group 20–59 years of age, there were no significant changes in the overall cohort or for any sex, race, or ethnicity group (**Supplemental Table 9**).

### ASMMI

For the earlier data cycles of 1999–2006, ASMMI did not significantly change overall or for any age, sex, race, or ethnicity group (**Figure 5, Supplemental Table 10**). In the 2011–2018 data cycles among participants 20–59 years of age, there was a significant increase in ASMMI for male survivors, from 8.62 kg/m^2^ (95% CI: 8.39–8.85) in 2011–2012 to 9.03 kg/m^2^ (95% CI: 8.87–9.20) in 2017–2018, with an average increase of 0.139 kg/m^2^ (P<.001). There was also a significant average increase of 0.13 kg/m^2^ per 2-year data cycle among non-Hispanic White survivors (P = .008) (**Supplemental Table 11**).

**Figure 5.**
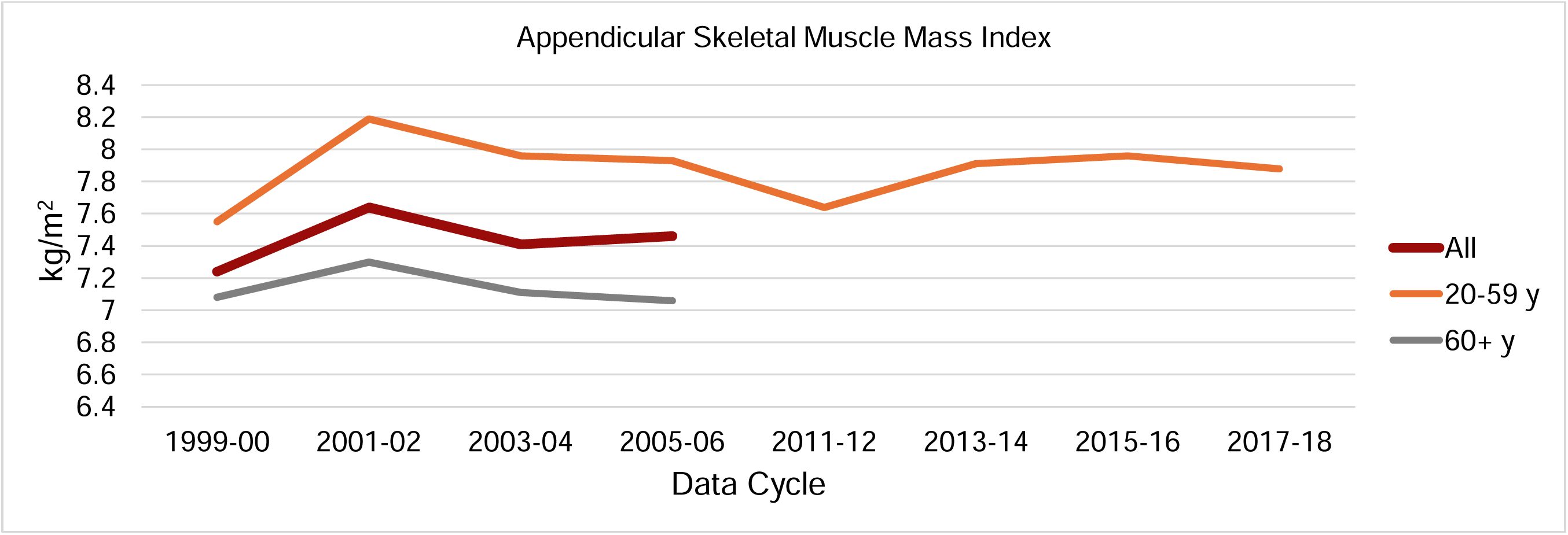
Adjusted^a^ appendicular skeletal muscle mass index averages: estimates overall and by age group^b^ in NHANES 1999 to 2018. ^a^ Estimates were adjusted for age (continuous), race and ethnicity, and sex. ^b^ All age group (includes all cancer survivors in the analysis, aged ≥20 years, 1999–2006): P = .34 and β = .040 per cycle. Age group 20–59 years, 1999–2006: P = .80 and β = .053; 2011–2018: P = .37 and β = .082. Age group 60+ years, 1999–2006: P = .39 and β = −.015.

### Sensitivity Analyses

Additionally adjusting for smoking status and physical activity levels showed trends that were similar to the main analyses, with BMI and WC increasing and FMI and ASMMI remaining unchanged over time (**Supplemental Tables 12 and 13**). LM showed an increasing trend during the 1999–2006 cycles. Among individuals who had DEXA data, none of the BMI trends were significant (i.e., for all ages in 1999–2006 cycles, age ≥60 years in 1999–2006, and age 20–59 years in 1999–2018 cycles) (**Supplemental Table 14**). However, WC increased for all ages in the 1999–2006 cycles (beta = 1.32, P=.02; **Supplemental Table 15**). Similar increasing trends in WC were observed among participants aged 60 years or older (1999–2006 cycles) and aged 20–59 years (1999– 2018 cycles) although these trends were not statistically significant. Additional adjustments for height, cancer type, or years since cancer diagnosis did not materially change the regression results (data not shown).

## Discussion

This longitudinal study reports the results of analyses spanning 19 years from a nationally representative sample of US cancer survivors, providing national estimates of body size and body composition measures, including BMI, WC, FMI, LM, and ASMMI. We observed overall significant increases in BMI and WC from 1999–2018, particularly among Hispanic and non-Hispanic White cancer survivors. Among individuals with DEXA data available, WC, but not BMI, showed an increasing trend, and LM increased significantly from 1999–2006. An increase in ASMMI was observed only among male and non-Hispanic White cancer survivors in the more recent cycles.

Our findings on BMI and WC are consistent with prior studies in the general US population using NHANES 2011–2018 data, including among individuals with or without cancer. One study showed significant increases in BMI and WC overall and for Hispanic, non-Hispanic White, and non-Hispanic Asian individuals.^2^ Similarly, a study of cancer survivors from the National Health Interview Survey reported an increase in self-reported BMI from 1997 to 2014.^3^ The current study observed analogous increases in both BMI and WC measures during the 1999 to 2018 time frame overall among cancer survivors and specifically in Hispanic and non-Hispanic White groups, but not the non-Hispanic Black group. Our findings suggest that obesity continues as a threat to the health of cancer survivors overall and within specific racial and ethnic groups.^8,28,29^

The increasing trends in LM and ASMMI over time are positive signs, as increased muscle mass is linked to a lower risk of sarcopenia, which is associated with better cancer survival.^15,16^ Increasing LM and appendicular muscle could be a result of exercise, particularly resistance training, which promotes muscle growth.^30^

We observed some discrepancies between the trends of anthropometric and DEXA-assessed body composition indices. The reason for the discrepancies may be multifactorial. BMI incorporates several components, including water mass, bone mass, organ mass, and height, in addition to adipose and muscle tissue mass. Based on our sensitivity analysis, height did not change over time. However, DEXA measurement errors in body fat and lean muscle may be greater in individuals with a higher vs lower BMI^31^ and may include a race and ethnicity differential error, as the computational algorithm was developed with less consideration of racial and ethnic diversity.^32^ Excess body water retention, a result of inflammation and lymphedema caused by lymph node resection, can contribute to a higher BMI among cancer survivors, but whether this factor increases over time is unclear. When we restricted the sample to individuals with DEXA data only, the BMI trend became more consistent with FMI, suggesting that the discrepancies may be due in part to the lack of DEXA data for individuals aged 60 years or older. Nevertheless, the increasing trend for WC in the restricted sample highlights the need for further research on abdominal obesity.

Aside from trend analyses, our data indicated that measures of BMI, WC, and FMI were elevated relative to obesity cutoffs. The 2017–2018 mean BMI estimate among cancer survivors reached 30.0 kg/m^2^, which is the World Health Organization–defined cutoff for obesity. Similarly, WC in 2017–2018 estimates of 100.9 cm for female cancer survivors and 106.2 cm for male cancer survivors surpassed the established thresholds of central obesity (i.e., ≥88 cm for female and ≥102 cm for male).^2^ The mean values of FMI also exceeded obesity cutoffs (≥ 6.6 kg/m^2^ in male and ≥ 9.5 kg/m^2^ in female),^33^ with male survivors at 8.40 kg/m^2^ and female survivors at 12.8 kg/m^2^ in 2006–2007. Given the association of high fat mass with inadequate exercise and unhealthy dietary patterns,^30,34,35^ the observed high BMI, WC, and FMI values could imply poor adherence to a healthy lifestyle among cancer survivors.^36^

The current study is the first analysis of body composition trends in cancer survivors. A strength of this study is our use of DEXA data, which provide precise measurements of fat and muscle mass that are key indicators linked to mortality, offering insights that anthropometric measures cannot differentiate. In addition, NHANES is a nationally representative sample, allowing for sample-weighted results that are generalizable to the US adult cancer survivor population. Our study also provides estimates adjusted by race and ethnicity, sex, and age to reduce confounding effects. Moreover, we were able to consider the confounding effects of physical activity and smoking on body fatness and composition trends.

This study has several limitations. From 2011–2018, NHANES DEXA data were restricted to individuals 20–59 years of age, preventing a full analysis of body composition in older (≥60 years) cancer survivors, a demographic group with typically higher cancer prevalence. Nevertheless, the data for individuals 20–59 years of age have clinical implications because incidences of geriatric-associated cancers have been increasing among young and middle-aged adults in recent decades.^37^ We were unable to present data for non-Hispanic Asian cancer survivors and other race or ethnicity groups in our race/ethnicity-specific analyses due to their small sample sizes. Additionally, NHANES included only non-institutionalized individuals and longer-term survivors. In the 2017–2018 cycle, the median time from the first cancer diagnosis to the interview was 7 years (interquartile range, 3–15 years). The sample may have underrepresented individuals with advanced cancer or undergoing treatment. Finally, causality cannot be inferred as the study design is observational.

## Conclusion

In this nationally representative longitudinal study, we examined trends in body fatness and changes in body composition among US cancer survivors from 1999 to 2018, leveraging DEXA scan data for measurements of fat and muscle mass. Our findings showed a significant increase across the 19-year period in BMI and WC, key risk factors for obesity-related mortality in this population. The continued upward trend in obesity measures and elevated FMI levels highlight the need for targeted interventions to address obesity-related health risks among cancer survivors. Adhering to healthy lifestyle guidelines more closely after a cancer diagnosis may help reduce body fat and increase muscle mass. Continuous body composition data collection for all adult cancer survivors is warranted to confirm the observed increased trends in LM and ASMMI.

## Data availability statement

NHANES data are publicly available at https://wwwn.cdc.gov/nchs/nhanes/Default.aspx

## Author contributions

Benjamin Brockbank, BS (Conceptualization; Formal analysis; Investigation; Methodology; Visualization; Writing – original draft; Writing – review & editing); Rand Talal Akasheh, PhD (Investigation; Methodology; Writing – original draft; Writing – review & editing); Yevgeniya Gokun, MS (Data curation; Formal analysis; Methodology; Validation; Writing – review & editing); Faiza Kalam, PhD (Conceptualization; Methodology; Writing – review & editing); Cuthbert Mario Mahenge, MD, MPH (Data curation; Project administration; Writing – review & editing); Electra D Paskett, PhD (Funding acquisition; Supervision; Writing – review & editing); Ting-Yuan David Cheng, Ph.D. (Conceptualization; Funding acquisition; Investigation; Methodology; Resources; Supervision; Writing – original draft; Writing – review & editing).

## Ethics approval

The study was deemed to be non-human subject research by the Institutional Review Board at the Ohio State University as the data were de-identified.

