## Supplemental Figure 1 for "Body Size and Composition Trends in U.S. Adults with a History of Cancer: NHANES 1999–2018"

**Supplemental Fig 1. Flowchart of sample selection.** Abbreviations: ASMMI, appendicular skeletal muscle mass index; BMI, body mass index; DEXA, dual-energy X-ray absorptiometry; FMI, fat mass index; NHANES, National Health and Nutrition Examination Survey; WC, waist circumference.

NHANES 1999-2006, 2011-2018 (n=80,630)

Adults aged 20 years or older (n=42,928)

Self-reported cancer history (n=3970)

Non-pregnant adults (n=3906)

Available BMI (n=3544)

Available WC

(n=3354)

Available DEXA (n=1951)

Excluded individuals under 20 years of age (n=37,702)

Excluded individuals without a history of cancer (n=38,958)

Excluded pregnant individuals (n=64)

Aged 20-59 y

(n=945)

Aged ≥60 y

(n=2599)

Aged 20-59 y

(n=923)

Aged ≥60 y

(n=2431)

Aged ≥60 y

(n=955)

Aged 20-59 y

(n=812)

Aged ≥60 y

(n=955)

Aged 20-59 y

(n=820)

Aged ≥60 y

(n=936)

Aged 20-59 y

(n=805)

Available FMI (n=1767)

Available LM

(n=1775)

Available ASMMI

(n=1741)

Available body measurements (n=3568)
