## Supplemental Tables 1-15 for "Body Size and Composition Trends in U.S. Adults with a History of Cancer: NHANES 1999–2018"

**Supplemental Table 1. Population characteristics for adult cancer survivors aged 20 years or older with body measurements available^a^ in NHANES, 1999–2006 and 2011–2018**

| Variables | **Totals** | | **1999-2000** | | **2001-2002** | | **2003-2004** | |
| --- | --- | --- | --- | --- | --- | --- | --- | --- |
|  | Count | Weighted percentage (95% CI) | Count | Weighted percentage (95% CI) | Count | Weighted percentage (95% CI) | Count | Weighted percentage (95% CI) |
| **Overall** | 3568 | / | 324 | / | 428 | / | 421 | / |
| **Age (years)** |  |  |  |  |  |  |  |  |
| 20–59 | 947 | 37.5 (35.1 to 40.0) | 74 | 36.9 (27.0 to 45.3) | 119 | 44.4 (39.7 to 49.0) | 99 | 39.6 (32.2 to 47.0) |
| ≥60 | 2621 | 62.5 (60.0 to 64.9) | 250 | 63.1 (54.7 to 73.0) | 309 | 55.6 (51.0 to 60.3) | 322 | 60.4 (53.0 to 67.8) |
| **Sex** |  |  |  |  |  |  |  |  |
| Female | 1870 | 57.3 (55.4 to 59.3) | 160 | 57.3 (52.5 to 62.0) | 218 | 59.8 (55.2 to 64.4) | 217 | 55.9 (52.0 to 59.8) |
| Male | 1698 | 42.7 (40.7 to 44.6) | 164 | 42.7 (38.0 to 47.5) | 210 | 40.2 (35.6 to 44.8) | 204 | 44.1 (40.2 to 48.0) |
| **Race/Ethnicity** |  |  |  |  |  |  |  |  |
| Hispanic | 436 | 4.8 (3.8 to 5.9) | 53 | 4.8 (1.3 to 8.3) | 38 | 4.9 (0.0 to 10.7) | 24 | 1.4 (0.2 to 2.6) |
| Non-Hispanic Black | 509 | 5.4 (4.4 to 6.3) | 39 | 5.8 (2.5 to 9.0) | 52 | 5.2 (2.9 to 7.4) | 48 | 5.9 (3.1 to 8.7) |
| Non-Hispanic White | 2439 | 86.2 (84.5 to 88.0) | 225 | 87.1 (81.8 to 92.3) | 334 | 88.9 (83.1 to 94.8) | 340 | 90.6 (86.7 to 94.5) |
| Other | 184 | 3.6 (2.7 to 4.5) | 7 | 2.4 (0.0 to 5.3) | 4 | 1.0 (0.0 to 2.3) | 9 | 2.0 (0.9 to 3.4) |
| **Smoking Status^b^** |  |  |  |  |  |  |  |  |
| Current | 546 | 16.9 (15.1 to 18.6) | 55 | 24.0 (17.3 to 30.7) | 56 | 15.9 (11.6 to 20.3) | 58 | 17.5 (12.8 to 22.3) |
| Former | 1448 | 38.9 (36.5 to 41.0) | 137 | 37.8 (31.0 to 33.6) | 199 | 42.1 (37.7 to 46.5) | 195 | 44.6 (38.2 to 51.1) |
| Never | 1572 | 44.4 (42.2 to 46.5) | 132 | 38.2 (31.8 to 44.6) | 173 | 42.0 (35.1 to 48.8) | 167 | 37.8 (32.6 to 43.1) |
| **Physical Activity^b^** |  |  |  |  |  |  |  |  |
| Inactive | 267 | 9.6 (8.1 to 11.1) | 41 | 18.8 (13.2 to 24.5) | 52 | 15.9 (9.9 to 21.8) | 50 | 14.7 (8.9 to 20.5) |
| Exceeding guidelines | 1155 | 50.8 (48.0 to 53.5) | 78 | 34.3 (25.7 to 42.9) | 102 | 35.3 (27.1 to 43.5) | 101 | 35.7 (30.4 to 41.1) |
| Meeting guidelines | 535 | 22.0 (19.7 to 24.3) | 47 | 22.1 (16.6 to 27.6) | 87 | 29.1 (23.5 to 34.7) | 80 | 25.7 (20.5 to 30.8) |
| Not meeting guidelines | 463 | 17.7 (16.0 to 19.4) | 48 | 24.7 (19.6 to 29.9) | 59 | 19.7 (14.6 to 24.8) | 78 | 23.9 (19.2 to 28.5) |

| **2005-2006** | | **2011-2012** | | **2013-2014** | | **2015-2016** | | **2017-2018** | |
| --- | --- | --- | --- | --- | --- | --- | --- | --- | --- |
| Count | Weighted percentage (95%CI) | Count | Weighted percentage (95%CI) | Count | Weighted percentage (95%CI) | Count | Weighted percentage (95%CI) | Count | Weighted percentage (95%CI) |
| 377 | / | 449 | / | 516 | / | 518 | / | 535 | / |
| 106 | 43.4 (35.6 to 51.2) | 126 | 40.0 (32.8 to 47.1) | 156 | 37.7 (31.8 to 43.7) | 133 | 32.3 (26.5 to 38.1) | 134 | 30.4 (23.7 to 37.0) |
| 271 | 56.6 (48.8 to 64.4) | 323 | 60.0 (52.9 to 67.2) | 360 | 62.3 (56.3 to 68.2) | 385 | 67.7 (61.9 to 73.5) | 401 | 69.7 (63.0 to 76.3) |
| 208 | 64.1 (58.3 to 69.8) | 229 | 57.2 (50.5 to 63.8) | 290 | 55.7 (51.3 to 60.0) | 264 | 54.7 (48.3 to 61.1) | 284 | 56.8 (52.0 to 61.5) |
| 169 | 35.9 (30.2 to 41.7) | 220 | 42.8 (36.2 to 49.5) | 226 | 44.3 (40.0 to 48.7) | 254 | 45.3 (38.9 to 51.7) | 251 | 43.2 (38.5 to 48.0) |
| 25 | 3.1 (1.4 to 4.7) | 56 | 5.3 (2.5 to 8.1) | 67 | 5.2 (3.1 to 7.3) | 94 | 5.0 (2.6 to 7.5) | 79 | 7.3 (4.0 to 10.5) |
| 53 | 5.2 (2.6 to 7.7) | 90 | 5.7 (2.8 to 8.7) | 69 | 4.7 (2.5 to 7.0) | 68 | 4.9 (2.4 to 7.4) | 90 | 5.8 (3.6 to 8.0) |
| 291 | 88.0 (84.9 to 91.0) | 270 | 86.1 (81.1 to 91.0) | 354 | 87.0 (83.1 to 90.9) | 316 | 83.9 (79.0 to 88.9) | 309 | 81.8 (76.0 to 87.6) |
| 8 | 3.8 (1.2 to 6.4) | 33 | 2.9 (1.8 to 4.1) | 26 | 3.0 (1.1 to 4.9) | 40 | 6.2 (2.5 to 9.9) | 57 | 5.2 (2.3 to 8.1) |
| 60 | 19.7 (13.9 to 25.5) | 64 | 14.5 (8.8 to 20.2) | 89 | 18.7 (15.2 to 22.3) | 85 | 15.1 (9.5 to 20.8) | 79 | 13.8 (10.8 to 16.8) |
| 152 | 38.0 (32.0 to 44.0) | 177 | 38.1 (32.3 to 43.8) | 193 | 37.0 (30.3 to 43.7) | 203 | 39.4 (33.3 to 45.5) | 192 | 35.3 (28.9 to 41.6) |
| 165 | 42.3 (36.2 to 48.4) | 208 | 47.4 (41.8 to 53.0) | 234 | 44.2 (37.6 to 50.9) | 229 | 45.4 (41.2 to 49.7) | 264 | 51.0 (44.0 to 58.0) |
| 39 | 13.0 (9.2 to 16.8) | 16 | 4.4 (1.5 to 7.2) | 24 | 5.8 (1.3 to 10.3) | 28 | 8.7 (4.9 to 12.4) | 17 | 3.4 (1.5 to 5.3) |
| 109 | 41.9 (34.1 to 49.6) | 151 | 52.2 (45.3 to 59.2) | 190 | 57.7 (52.9 to 62.5) | 206 | 61.7 (52.3 to 71.2) | 218 | 66.5 (59.3 to 73.8) |
| 65 | 23.5 (18.6 to 28.5) | 61 | 23.0 (16.5 to 29.4) | 72 | 19.9 (14.6 to 25.2) | 62 | 16.9 (9.3 to 24.5) | 61 | 20.1 (13.6 to 26.7) |
| 64 | 21.6 (14.8 to 28.3) | 50 | 20.5 (16.5 to 24.4) | 64 | 16.6 (11.0 to 22.2) | 59 | 12.7 (9.5 to 15.8) | 41 | 9.9 (6.1 to 13.8) |

^a^ Body measurement inclusion was defined as participant data available on either body mass index or waist circumference

^b^ Physical activity and smoking status totals do not add to 100% because some participants had missing or unavailable data for these variables

**Supplemental Table 2. Participants available on select NHANES variables by age, sex, race and ethnicity, and survey cycle: 1999–2006 and 2011–2018**

|  | Participants with BMI data, n (%) | Participants with WC data, n (%) | Participants with DEXA data^a^, n (%) |
| --- | --- | --- | --- |
| Total | 3544 (100) | 3354 (100) | 1951 (100) |
| Age (years) |  |  |  |
| 20–59 | 945 (27) | 923 (28) | 961 (49) |
| ≥60 | 2599 (73) | 2431 (82) | 990 (51) |
| Sex |  |  |  |
| Female | 1859 (52) | 1755 (52) | 1107 (57) |
| Male | 1685 (48) | 1599 (48) | 844 (43) |
| Race/Ethnicity |  |  |  |
| Hispanic | 433 (12) | 409 (12) | 236 (12) |
| NH Black | 506 (14) | 474 (14) | 262 (13) |
| NH White | 2421 (68) | 2298 (69) | 1366 (70) |
| Other | 184 (5) | 173 (5) | 87 (4) |
| Survey Cycle |  |  |  |
| 1999–2006 | 1527 (43) | 1489 (44) | 1391 (71) |
| 2011–2018 | 2017 (57) | 1865 (56) | 560 (29) |

^a^ Inclusion for participants with DEXA data available in NHANES

Abbreviations: BMI, body mass index; DEXA, dual-energy X-ray absorptiometry; NH, non-Hispanic; NHANES, National Health and Nutrition Examination Survey; WC, waist circumference

**Supplemental Table 3. Frequency (and relative %) of first cancer type for cancer survivors aged 20 years or older with body measurements available in NHANES sample**

| **By Cancer Type** | **Total, n (%)** | **Female, n (%)** | **Male, n (%)** |
| --- | --- | --- | --- |
| Obesity-associated^a^ | 1140 (32) | 918 (49) | 222 (13) |
| Non-obesity associated | 2428 (68) | 952 (51) | 1476 (87) |
| **By Cancer Location** |  |  |  |
| Breast | 550 | 550 | 0 |
| Prostate | 542 | 0 | 542 |
| Skin (non-melanoma) | 520 | 225 | 295 |
| Skin (unknown type) | 280 | 123 | 157 |
| Melanoma | 217 | 97 | 120 |
| Colon | 218 | 102 | 116 |
| Cervix | 202 | 202 | 0 |
| Uterus | 141 | 141 | 0 |
| Lung | 99 | 35 | 64 |
| Bladder | 87 | 19 | 68 |
| Ovarian | 72 | 72 | 0 |
| Lymphoma | 70 | 35 | 35 |
| Kidney | 70 | 23 | 47 |
| Thyroid | 67 | 57 | 10 |
| Other | 433 | 189 | 244 |

^a^ Obesity-associated cancers per Centers for Disease Control and Prevention–posted risk factors include adenocarcinoma of esophagus, breast, colon and rectum, uterus, gallbladder, upper stomach, kidneys, liver, ovaries, pancreas, thyroid, meningioma and multiple myeloma. Table shows 14 most prevalent cancer types within the study population, with all other cancers in “other” category. NHANES-provided variable based on the survey question for type of first cancer: “What kind of cancer was it?”

**Supplemental Table 4. Age-, race/ethnicity-, and sex-adjusted^a^ mean body mass index (BMI) levels among cancer survivors 20 years of age or older in the United States, 1999–2018 (n=3544)**

| **Variable** | BMI (kg/m^2^), mean (95% CI) | | | | | | | | **Beta** | **P value** |
| --- | --- | --- | --- | --- | --- | --- | --- | --- | --- | --- |
|  | **1999-2000** | **2001-2002** | **2003-2004** | **2005-2006** | **2011-2012** | **2013-2014** | **2015-2016** | **2017-2018** |  |  |
| Overall | 27.1 (25.9 to 28.3) | 29.2 (28.3 to 30.0) | 28.5 (27.9 to 29.1) | 28.2 (27.4 to 28.9) | 29.2 (28.4 to 30.1) | 29.5 (28.8 to 30.3) | 29.9 (29.1 to 30.6) | 30.0 (29.4 to 30.6) | 0.233 | **<0.001** |
| **Age (years)** |  |  |  |  |  |  |  |  |  |  |
| 20–59 | 27.0 (25.5 to 28.6) | 30.5 (29.1 to 31.9) | 29.8 (28.3 to 31.4) | 29.4 (27.9 to 30.9) | 30.5 (28.7 to 32.3) | 30.8 (29.4 to 32.3) | 30.8 (29.6 to 31.9) | 30.4 (28.7 to 32.1) | 0.204 | **0.002** |
| ≥60´ | 27.0 (25.8 to 28.2) | 28.1 (27.0 to 29.3) | 27.6 (26.9 to 28.3) | 27.3 (26.5 to 28.2) | 28.2 (27.6 to 28.9) | 28.5 (27.7 to 29.2) | 29.2 (28.2 to 30.1) | 29.4 (28.7 to 30.0) | 0.232 | **<0.001** |
| **Sex** |  |  |  |  |  |  |  |  |  |  |
| Female | 27.1 (25.5 to 28.7) | 30.5 (29.3 to 31.7) | 28.8 (27.9 to 29.7) | 28.7 (27.5 to 29.9) | 30.2 (28.7 to 31.7) | 30.6 (29.7 to 31.4) | 29.9 (28.9 to 31.0) | 30.3 (29.3 to 31.3) | 0.260 | **<0.001** |
| Male | 27.6 (26.1 to 28.1) | 27.2 (26.4 to 28.7) | 27.8 (27.0 to 28.6) | 27.4 (26.5 to 28.4) | 27.8 (26.8 to 28.8) | 28.1 (26.8 to 29.3) | 29.6 (28.9 to 30.4) | 29.4 (28.4 to 30.5) | 0.212 | **<0.001** |
| **Race/Ethnicity** |  |  |  |  |  |  |  |  |  |  |
| Hispanic | 27.6 (25.9 to 29.3) | 29.8 (28.1 to 31.5) | 28.0 (24.3 to 31.7) | 27.8 (25.5 to 30.1) | 30.2 (29.1 to 31.3) | 29.8 (27.9 to 31.6) | 31.5 (30.1 to 32.9) | 30.6 (28.8 to 32.5) | 0.311 | **0.001** |
| Non-Hispanic Black | 27.6 (25.7 to 29.5) | 29.3 (26.9 to 31.7) | 29.9 (27.5 to 32.3) | 31.0 (28.8 to 33.2) | 31.0 (29.1 to 33.0) | 29.4 (27.7 to 31.1) | 30.1 (28.5 to 31.7) | 29.6 (28.7 to 30.6) | 0.090 | 0.288 |
| Non-Hispanic White | 26.4 (25.3 to 27.5) | 28.6 (27.7 to 29.4) | 27.7 (27.2 to 28.1) | 27.5 (26.8 to 28.1) | 28.6 (27.6 to 29.6) | 28.9 (28.1 to 29.7) | 29.2 (28.4 to 30.1) | 29.4 (28.9 to 30.0) | 0.245 | **<0.001** |
| Other Race/Ethnicity | 29.6 (25.2 to 34.0) | 26.7 (23.7 to 29.8) | 31.2 (25.8 to 36.6) | 25.4 (22.5 to 28.4) | 26.0 (24.0 to 28.0) | 30.3 (26.9 to 33.6) | 28.8 (27.0 to 30.6) | 27.9 (26.5 to 29.3) | 0.048 | 0.159 |

^a^ Overall BMI was adjusted by age (continuous), sex, and race/ethnicity. Race/ethnicity and sex estimates were not adjusted for race/ethnicity and sex, respectively.

**Supplemental Table 5. Age- race/ethnicity- and sex-adjusted^a^ mean waist circumference among United States cancer survivors ages 20 years or older, 1999–2018 (n=3354)**

| **Variable** | Adjusted Waist Circumference (cm), mean (95% confidence interval) | | | | | | | | **Beta** | **P value** |
| --- | --- | --- | --- | --- | --- | --- | --- | --- | --- | --- |
|  | **1999-2000** | **2001-2002** | **2003-2004** | **2005-2006** | **2011-2012** | **2013-2014** | **2015-2016** | **2017-2018** |  |  |
| Overall | 96.1 (93.2 to 99.1) | 101.6 (100.2 to 103.1) | 101.3 (99.5 to 103.0) | 99.5 (97.7 to 101.3) | 102.8 (100.7 to 104.8) | 103.1 (101.2 to 105.1) | 104.0 (102.2 to 105.8) | 103.8 (102.2 to 105.5) | 0.599 | **<0.001** |
| **Age (years)** |  |  |  |  |  |  |  |  |  |  |
| 20–59 | 93.9 (90.2 to 97.7) | 102.3 (98.9 to 105.8) | 102.4 (98.9 to 105.9) | 99.6 (96.3 to 102.0) | 102.8 (98.6 to 106.9) | 104.1 (100.7 to 107.6) | 103.6 (100.8 to 106.4) | 102.4 (98.0 to 106.7) | 0.523 | **<0.001** |
| ≥60 | 96.3 (93.3 to 99.2) | 100.0 (98.4 to 101.6) | 99.5 (96.7 to 101.6) | 98.5 (96.7 to 100.3) | 101.4 (99.5 to 103.3) | 101.1 (99.4 to 102.8) | 103.0 (100.6 to 105.3) | 102.9 (101.2 to 104.7) | 0.582 | **<0.001** |
| **Sex** |  |  |  |  |  |  |  |  |  |  |
| Female | 91.6 (87.5 to 95.8) | 100.2 (97.7 to 102.7) | 97.7 (95.3 to 100.1) | 96.4 (93.6 to 99.1) | 100.9 (97.2 to 104.6) | 101.5 (99.4 to 103.5) | 100.0 (97.8 to 102.2) | 100.9 (98.3 to 103.5) | 0.620 | **<0.001** |
| Male | 100.6 (98.0 to 103.2) | 101.8 (99.3 to 104.4) | 104.2 (101.9 to 106.5) | 102.4 (99.7 to 105.1) | 103.8 (100.9 to 106.6) | 104.0 (100.8 to 107.1) | 107.6 (105.5 to 109.8) | 106.2 (103.4 to 109.0) | 0.575 | **<0.001** |
| **Race/Ethnicity** |  |  |  |  |  |  |  |  |  |  |
| Hispanic | 94.4 (90.0 to 98.9) | 102.3 (96.7 to 107.8) | 93.2 (88.6 to 97.8) | 97.1 (93.0 to 101.3) | 102.3 (99.6 to 104.9) | 102.6 (98.0 to 107.2) | 106.1 (103.2 to 108.9) | 105.1 (101.5 to 108.7) | 0.961 | **<0.001** |
| Non-Hispanic Black | 96.0 (90.8 to 101.3) | 99.4 (95.3 to 103.5) | 102.4 (96.7 to 108.0) | 103.2 (98.8 to 107.7) | 104.6 (101.6 to 107.6) | 100.6 (96.5 to 104.6) | 103.4 (99.9 to 107.0) | 102.2 (99.9 to 104.5) | 0.400 | 0.155 |
| Non-Hispanic White | 95.7 (93.0 to 98.4) | 101.4 (100.1 to 102.8) | 100.7 (99.4 to 102.1) | 98.9 (97.2 to 100.6) | 102.5 (99.9 to 105.1) | 102.8 (101.0 to 104.7) | 103.6 (101.6 to 105.6) | 103.5 (101.7 to 105.3) | 0.605 | **<0.001** |
| Other Race/Ethnicity | 100.9 (88.7 to 113.1) | 94.7 (87.7 to 101.8) | 106.1 (94.1 to 118.1) | 97.8 (90.9 to 104.7) | 96.2 (90.7 to 101.8) | 105.2 (96.7 to 113.7) | 102.9 (97.6 to 108.1) | 99.9 (95.3 to 104.4) | 0.204 | 0.381 |

^a^ Overall waist circumference was adjusted by age (continuous), sex, and race/ethnicity. Race/ethnicity estimates were adjusted for age and sex. Sex estimates were adjusted for age and race/ethnicity.

**Supplemental Table 6. Age- race/ethnicity- and sex-adjusted^a^ mean fat mass index in United States cancer survivors ages 20 years or older, 1999–2006 (n=822)**

| **Variable** | Fat Mass Index (kg/m^2^), mean (95% confidence interval) | | | | **Beta** | **P value** |
| --- | --- | --- | --- | --- | --- | --- |
|  | **1999-2000** | **2001-2002** | **2003-2004** | **2005-2006** |  |  |
| Overall | 10.1 (9.32 to 10.9) | 11.4 (10.8 to 12.0) | 10.9 (10.3 to 11.5) | 10.9 (10.1 to 11.6) | 0.172 | 0.225 |
| **Age (years)** |  |  |  |  |  |  |
| 20–59 | 9.57 (8.47 to 10.7) | 11.7 (10.6 to 12.8) | 11.3 (10.1 to 12.4) | 10.8 (9.70 to 11.9) | 0.191 | 0.381 |
| ≥60 | 10.2 (9.40 to 10.9) | 10.8 (10.1 to 11.5) | 10.4 (9.89 to 10.9) | 10.4 (9.24 to 11.5) | 0.044 | 0.779 |
| **Sex** |  |  |  |  |  |  |
| Female | 11.4 (10.3 to 12.5) | 13.7 (12.8 to 14.5) | 12.5 (11.7 to 13.4) | 12.8 (11.9 to 13.8) | 0.255 | 0.180 |
| Male | 8.65 (7.86 to 9.43) | 8.58 (8.03 to 9.13) | 8.93 (8.34 to 9.53) | 8.40 (7.46 to 9.33) | 0.015 | 0.916 |
| **Race/Ethnicity^b^** |  |  |  |  |  |  |
| Hispanic | 10.1 (9.44 to 10.8) | 10.9 (10.0 to 11.8) | 10.3 (10.1 to 10.6) | 9.56 (9.08 to 10.0) | -0.226 | **0.049** |
| Non-Hispanic Black | 9.78 (8.60 to 11.0) | 11.1 (9.63 to 12.5) | 11.1 (9.79 to 12.3) | 11.6 (10.2 to 12.9) | 0.530 | 0.053 |
| Non-Hispanic White | 9.57 (8.92 to 10.2) | 10.9 (10.4 to 11.3) | 10.3 (9.99 to 10.6) | 10.4 (9.8 to 11.1) | 0.193 | 0.167 |

^a^ Overall fat mass index was adjusted by age (continuous), sex and race/ethnicity. Race/ethnicity and sex estimates were not adjusted for race/ethnicity and sex respectively.

^b^ “Non-Hispanic Other” group was not included due to small sample size.

**Supplemental Table 7. Age- race/ethnicity- and sex-adjusted^a^ mean fat mass index among United States cancer survivors ages 20–59 years, 2011–2018**

| **Variable** | Fat Mass Index (kg/m^2^), mean (95% confidence interval) | | | | **Beta** | **P value** |
| --- | --- | --- | --- | --- | --- | --- |
|  | **2011-2012** | **2013-2014** | **2015-2016** | **2017-2018** |  |  |
| Overall | 10.9 (9.42 to 12.4) | 11.4 (9.94 to 12.8) | 11.3 (10.2 to 12.3) | 10.5 (8.81 to 12.1) | -0.121 | 0.687 |
| **Sex** |  |  |  |  |  |  |
| Female | 13.1 (10.6 to 15.6) | 14.1 (12.0 to 16.1) | 13.4 (12.4 to 14.5) | 12.8 (11.5 to 14.1) | -0.137 | 0.697 |
| Male | 8.59 (7.88 to 9.30) | 8.31 (7.94 to 8.68) | 8.92 (8.56 to 9.29) | 7.85 (7.50 to 8.20) | -0.121 | 0.198 |
| **Race/Ethnicity^b^** |  |  |  |  |  |  |
| Hispanic | NA | NA | NA | NA | NA | NA |
| Non-Hispanic Black | NA | NA | NA | NA | NA | NA |
| Non-Hispanic White | 10.1 (8.74 to 11.5) | 10.7 (10.4 to 11.0) | 10.7 (10.4 to 11.0) | 9.6 (9.31 to 9.86) | -0.096 | 0.625 |

NA, not available, was a result of inability to perform statistical analysis due to small sample size.

^a^ Overall fat mass index was adjusted by age (continuous), sex, and race/ethnicity. Race/ethnicity estimates were adjusted for age and sex. Sex estimates were adjusted for age and race/ethnicity.

^b^ “Non-Hispanic other” group was not included due to small sample size.

**Supplemental Table 8. Age- race/ethnicity- and sex-adjusted^a^ mean total lean mass among United States cancer survivors ages 20–59 years, 1999–2006**

| **Variables** | Total Lean Mass (kg), mean (95% confidence interval) | | | | **Beta** | **P value** |
| --- | --- | --- | --- | --- | --- | --- |
|  | **1999-2000** | **2001-2002** | **2003-2004** | **2005-2006** |  |  |
| Overall | 46.6 (45.0 to 48.2) | 49.3 (48.1 to 50.6) | 48.9 (47.6 to 50.2) | 49.5 (47.9 to 51.1) | 0.824 | **0.002** |
| **Age (years)** |  |  |  |  |  |  |
| 20–59 | 49.7 (47.7 to 51.7) | 54.2 (52.4 to 55.9) | 53.3 (51.1 to 55.5) | 53.2 (50.9 to 55.4) | 0.730 | 0.093 |
| ≥60 | 44.9 (42.8 to 47.1) | 46.1 (44.3 to 48.0) | 46.3 (44.6 to 47.9) | 45.8 (42.3 to 49.4) | 0.429 | 0.256 |
| **Sex** |  |  |  |  |  |  |
| Female | 39.6 (38.1 to 41.2) | 43.0 (41.5 to 44.4) | 41.3 (39.9 to 42.8) | 42.7 (40.8 to 44.5) | 0.658 | **0.027** |
| Male | 53.0 (50.7 to 55.4) | 54.8 (52.6 to 57.0) | 55.6 (53.4 to 57.9) | 55.7 (52.1 to 59.2) | 0.964 | **0.038** |
| **Race/Ethnicity^b^** |  |  |  |  |  |  |
| Hispanic | 44.8 (42.2 to 47.5) | 45.3 (43.0 to 47.6) | 46.6 (44.1 to 49.1) | 48.3 (45.0 to 51.6) | 1.174 | 0.057 |
| Non-Hispanic Black | 50.4 (48.0 to 52.9) | 51.0 (48.4 to 53.6) | 53.0 (50.5 to 55.6) | 53.1 (49.8 to 56.4) | 1.049 | 0.091 |
| Non-Hispanic White | 46.8 (45.8 to 47.8) | 49.7 (48.8 to 50.6) | 48.9 (47.9 to 49.9) | 49.7 (48.0 to 51.3) | 0.743 | **0.009** |

^a^ Overall lean mass was adjusted by age (continuous), sex, and race/ethnicity. Race/ethnicity and sex estimates were not adjusted for race/ethnicity and sex, respectively.

^b^ “Non-Hispanic other” group was excluded due to small sample size.

**Supplemental Table 9. Age- race/ethnicity- and sex-adjusted^a^ total lean mass among United States cancer survivors ages 20–59 years, 2011–2018**

| **Variable** | Total Lean Mass (kg), mean (95% confidence interval) | | | | **Beta** | **P value** |
| --- | --- | --- | --- | --- | --- | --- |
|  | **2011-2012** | **2013-2014** | **2015-2016** | **2017-2018** |  |  |
| Overall | 51.8 (48.4 to 55.2) | 52.9 (49.2 to 56.5) | 52.1 (49.7 to 54.6) | 51.5 (47.5 to 55.5) | -0.150 | 0.814 |
| **Sex** |  |  |  |  |  |  |
| Female | 43.8 (39.7 to 47.8) | 46.2 (42.2 to 50.2) | 44.1 (42.1 to 46.0) | 43.1 (41.0 to 45.2) | -0.414 | 0.503 |
| Male | 59.6 (57.9 to 61.3) | 58.4 (56.8 to 59.9) | 60.0 (58.2 to 61.8) | 60.1 (58.4 to 61.9) | 0.292 | 0.227 |
| **Race/Ethnicity^b^** |  |  |  |  |  |  |
| Hispanic | NA | NA | NA | NA | NA | NA |
| Non-Hispanic Black | NA | NA | NA | NA | NA | NA |
| Non-Hispanic White | 50.9 (48.4 to 53.5) | 51.7 (50.6 to 52.7) | 51.6 (50.5 to 52.7) | 50.9 (49.7 to 52.1) | 0.024 | 0.953 |

NA, not available, was a result of inability to perform statistical analysis due to small sample size.

^a^ Overall lean mass was adjusted by age (continuous), sex, and race/ethnicity. Race/ethnicity estimates were adjusted for age and sex. Sex estimates were adjusted for age and race/ethnicity.

^b^ “Non-Hispanic other” group was not included due to small sample size.

**Supplemental Table 10. Age- race/ethnicity- and sex-adjusted^a^ mean appendicular skeletal muscle mass index among United States cancer survivors ages 20 years or older, 1999–2006 (N=822)**

| **Variable** | Appendicular Skeletal Muscle Mass Index (kg/m^2^), mean (95% confidence interval) | | | | **Beta** | **P value** |
| --- | --- | --- | --- | --- | --- | --- |
|  | **1999-2000** | **2001-2002** | **2003-2004** | **2005-2006** |  |  |
| Overall | 7.24 (6.98 to 7.49) | 7.64 (7.39 to 7.89) | 7.41 (7.26 to 7.58) | 7.46 (7.23 to 7.69) | 0.040 | 0.338 |
| **Age (years)** |  |  |  |  |  |  |
| 20–59 | 7.55 (7.24 to 7.87) | 8.19 (7.91 to 8.47) | 7.96 (7.66 to 8.26) | 7.93 (7.62 to 8.23) | 0.053 | 0.803 |
| ≥60 | 7.08 (6.77 to 7.40) | 7.30 (6.96 to 7.64) | 7.11 (6.90 to 7.31) | 7.06 (6.60 to 7.53) | -0.015 | 0.394 |
| **Sex** |  |  |  |  |  |  |
| Female | 6.58 (6.29 to 6.86) | 7.10 (6.77 to 7.45) | 6.72 (6.50 to 6.93) | 6.84 (6.52 to 7.16) | 0.025 | 0.377 |
| Male | 7.85 (7.56 to 8.15) | 8.06 (7.80 to 8.32) | 8.02 (7.79 to 8.26) | 8.00 (7.65 to 8.36) | 0.048 | 0.395 |
| **Race/Ethnicity^b^** |  |  |  |  |  |  |
| Hispanic | 7.35 (7.05 to 7.66) | 7.15 (6.80 to 7.50) | 7.47 (7.36 to 7.59) | 7.33 (7.02 to 7.64) | 0.019 | 0.719 |
| Non-Hispanic Black | 8.02 (7.63 to 8.41) | 8.14 (7.64 to 8.64) | 8.21 (7.77 to 8.65) | 8.34 (7.81 to 8.88) | 0.102 | 0.309 |
| Non-Hispanic White | 6.91 (6.73 to 7.09) | 7.36 (7.11 to 7.61) | 7.08 (6.98 to 7.18) | 7.17 (6.95 to 7.39) | 0.040 | 0.363 |

^a^ Overall appendicular skeletal muscle mass index was adjusted by age (continuous), sex, and race/ethnicity. Race/ethnicity estimates were adjusted for age and sex. Sex estimates were adjusted for age and race/ethnicity.

^b^ “Non-Hispanic other” group was not included due to small sample size.

**Supplemental Table 11. Age- race/ethnicity- and sex-adjusted^a^ mean appendicular skeletal muscle mass index among United States cancer survivors ages 20–59 years, 2011–2018**

| **Variables** | Appendicular Skeletal Muscle Mass (kg/m^2^) mean (95% confidence interval) | | | | **Beta** | **P value** |
| --- | --- | --- | --- | --- | --- | --- |
|  | **2011-2012** | **2013-2014** | **2015-2016** | **2017-2018** |  |  |
| Overall | 7.64 (7.15 to 8.14) | 7.91 (7.43 to 8.39) | 7.96 (7.56 to 8.37) | 7.88 (7.29 to 8.48) | 0.082 | 0.367 |
| **Sex** |  |  |  |  |  |  |
| Female | 7.26 (6.66 to 7.87) | 7.17 (6.89 to 7.45) | 7.17 (6.89 to 7.45) | 6.89 (6.57 to 7.21) | -0.048 | 0.546 |
| Male | 8.62 (8.39 to 8.85) | 8.45 (8.31 to 8.59) | 8.75 (8.59 to 8.91) | 9.03 (8.87 to 9.20) | 0.139 | **<0.001** |
| **Race/Ethnicity^b^** |  |  |  |  |  |  |
| Hispanic | NA | NA | NA | NA | NA | NA |
| Non-Hispanic Black | NA | NA | NA | NA | NA | NA |
| Non-Hispanic White | 7.13 (6.80 to 7.47) | 7.45 (7.34 to 7.57) | 7.53 (7.42 to 7.65) | 7.50 (7.37 to 7.64) | 0.130 | **0.008** |

NA, not available, was a result of inability to perform statistical analysis due to small sample size.

^a^ Overall appendicular skeletal muscle mass index was adjusted by age (continuous), sex, and race/ethnicity. Race/ethnicity and sex estimates were not adjusted for race/ethnicity and sex, respectively.

^b^ “Non-Hispanic other” group was not included due to small sample size.

**Supplemental Table 12. Adjusted^a^ smoking status model-based means along with 95% confidence interval (CIs) for each outcome**

| **Data Cycle** | **BMI**  **Mean (95% CI)^b^** | **WC**  **Mean (95% CI)^c^** | **FMI**  **Mean (95% CI)^d^** | **LM**  **Mean (95% CI)^e^** | **ASMMI**  **Mean (95% CI)^f^** |
| --- | --- | --- | --- | --- | --- |
| 1999-2000 | 26.9 (25.7-28.1) | 95.8 (92.9-98.7) | 10.0 (9.2-10.8) | 46.3 (44.7-47.8) | 7.2 (6.9-7.4) |
| 2001-2002 | 28.8 (28.0-29.7) | 101.1 (99.6-102.5) | 11.2 (10.6-11.8) | 48.8 (47.6-50.0) | 7.5 (7.3-7.8) |
| 2003-2004 | 28.2 (27.5-28.8) | 100.8 (98.9-102.6) | 10.7 (10.1-11.3) | 48.5 (47.1-49.8) | 7.3 (7.2-7.5) |
| 2005-2006 | 27.9 (27.1-28.7) | 99.1 (97.3-100.9) | 10.7 (9.9-11.5) | 49.1 (47.5-50.7) | 7.4 (7.1-7.6) |
| 2011-2012 | 28.9 (28.1-29.7) | 102.4 (100.4-104.4) | 10.7 (9.2-12.2)^g^ | 51.4 (48.0-54.7)^g^ | 7.6 (7.1-8.1)^g^ |
| 2013-2014 | 29.3 (28.5-30.1) | 102.8 (100.8-104.8) | 11.3 (9.9-12.8)^g^ | 52.8 (49.2-56.4)^g^ | 7.9 (7.4-8.4)^g^ |
| 2015-2016 | 29.6 (28.8-30.4) | 103.6 (101.7-105.5) | 11.1 (10.0-12.3)^g^ | 51.7 (49.1-54.2)^g^ | 7.9 (7.5-8.3)^g^ |
| 2017-2018 | 29.7 (29.9-30.4) | 103.5 (101.9-105.1) | 10.3 (8.5-12.0)^g^ | 50.9 (47.0-54.8)^g^ | 7.8 (7.2-8.4)^g^ |

Abbreviations: ASMMI, appendicular skeletal muscle mass index; BMI, body mass index, FMI, fat mass index; LM, lean mass; WC waist circumference.

^a^ The model included NHANES cycles, sex (male, female), age (continuous), race/ethnicity category (non-Hispanic White, non-Hispanic African American/Black, Hispanic, and non-Hispanic other), and smoking status (current, former and never) as predictors.

^b^ Beta estimate for cycle was **.233 (P<.001)** in the BMI model.

^c^ Beta estimate for cycle was **.611 (P<.001)** in the WC model.

^d^ Beta estimate for cycle was .036 (P= .39) during 1999-2006 and .073 (P = .44) during 2011-2018 in the ASMMI model.

^e^ Beta estimate for cycle was .162 (P= .24) during 1999-2006 and -.137 (P = .65) during 2011-2018 in the FMI model.

^f^ Beta estimate for cycle was **.800 (P= .003)** during 1999-2006 and -.212 (P = .74) during 2011-2018 in the LM model.

^g^ Age 20–59 years only

**Supplemental Table 13. Adjusted^a^ physical activity model-based means along with 95% confidence interval (CIs) for each outcome**

| **Data Cycle** | **BMI**  **Mean (95% CI)^b^** | **WC**  **Mean (95% CI)^c^** | **FMI**  **Mean (95% CI)^d^** | **LM**  **Mean (95% CI)^e^** | **ASMMI**  **Mean (95% CI)^f^** |
| --- | --- | --- | --- | --- | --- |
| 1999-2000 | 27.8 (26.6-28.9) | 97.1 (94.2-100.0) | 10.5 (9.6-11.4) | 48.0 (45.9-50.1) | 7.4 (7.1-7.7) |
| 2001-2002 | 28.9 (28.1-29.8) | 100.5 (98.5-102.4) | 11.2 (10.5-12.0) | 49.6 (48.2-51.0) | 7.6 (7.4-7.9) |
| 2003-2004 | 29.1 (28.2-29.9) | 102.4 (100.4-104.3) | 11.2 (10.4-12.0) | 50.2 (48.7-51.7) | 7.6 (7.4-7.8) |
| 2005-2006 | 28.7 (27.7-29.7) | 100.5 (98.0-102.9) | 11.0 (10.1-11.8) | 50.5 (48.7-52.3) | 7.6 (7.3-7.8) |
| 2011-2012 | 29.7 (28.6-30.8) | 103.6 (101.0-106.1) | 11.0 (9.1-12.9)^g^ | 52.5 (48.1-56.9)^g^ | 7.9 (7.3-8.4)^g^ |
| 2013-2014 | 29.7 (28.8-30.7) | 103.4 (101.1-105.7) | 11.9 (10.3-13.4)^g^ | 54.2 (50.5-57.9)^g^ | 8.1 (7.7-8.6)^g^ |
| 2015-2016 | 30.3 (29.5-31.2) | 104.8 (102.8-106.8) | 11.5 (10.0-13.1)^g^ | 53.2 (50.5-55.8)^g^ | 8.2 (7.8-8.7)^g^ |
| 2017-2018 | 30.1 (29.2-31.0) | 103.7 (101.4-106.1) | 10.4 (8.5-12.3)^g^ | 51.5 (46.9-56.1)^g^ | 7.9 (7.1-8.8)^g^ |

Abbreviations: ASMMI, appendicular skeletal muscle mass index; BMI, body mass index, FMI, fat mass index; LM, lean mass; WC waist circumference.

^a^ The model included NHANES cycles, sex (male, female), age (continuous), race/ethnicity category (non-Hispanic White, non-Hispanic African American/Black, Hispanic, and non-Hispanic other), and leisure-time physical activity (inactive, insufficiently active, active, and highly active) as predictors.

^b^ Beta estimate for cycle was **.219 (P<.001)** in the BMI model (n = 2408).

^c^ Beta estimate for cycle was **.588 (P<.001)** in the WC model (n = 2336).

^d^ Beta estimate for cycle was .037 (P = .42) during 1999-2006 (n = 956) and .039 (P = 0.73) during 2011-2018 (n = 311) in the FMI model.

^e^ Beta estimate for cycle was .140 (P = .34) during 1999-2006 (n = 956) and -.194 (P = 0.53) during 2011-2018 (n = 313) in the LM model.

^f^ Beta estimate for cycle was **.810 (P = .01) during 1999-2006** (n = 966) and -.366 (P = 0.63) during 2011-2018 (n = 319) in the ASMMI model.

^g^ Age 20–59 years only

**Supplemental Table 14. Adjusted^a^ model-based means along with 95% confidence interval (CIs) for BMI among cancer survivors with non-missing fat mass index**

| **Data Cycle** | **Among age 20–59 years^b^** | **Among ages ≥60 years^c^** | **Across All Ages^d^** |
| --- | --- | --- | --- |
| 1999-2000 | 27.2 (25.6-28.7) | 27.3 (26.0-28.7) | 27.6 (26.3-28.9) |
| 2001-2002 | 30.8 (29.3-32.2) | 28.5 (27.2-29.8) | 29.8 (28.8-30.7) |
| 2003-2004 | 29.9 (28.4-31.5) | 27.8 (27.0-28.6) | 28.9 (28.1-29.8) |
| 2005-2006 | 29.5 (27.9-31.0) | 28.0 (25.9-30.1) | 29.2 (28.0-30.3) |
| 2011-2012 | 30.0 (27.9-32.0) | NA | NA |
| 2013-2014 | 30.9 (29.0-32.8) | NA | NA |
| 2015-2016 | 30.8 (29.2-32.4) | NA | NA |
| 2017-2018 | 29.9 (27.9-31.8) | NA | NA |

NA, not available per NHANES survey age limitation for DEXA scans.

^a^ The model included NHANES cycles, sex (male, female), age (continuous), and race/ethnicity category (non-Hispanic African American/Black, non-Hispanic White, Hispanic, and non-Hispanic other).

^b^ Beta estimate for cycle was .156 (P= .12) in this model (n = 807).

^c^ Beta estimate for cycle was .155 (P= .57) in this model (n = 930). Data cycles of 2011-2018 were not included in this analysis because ≥60 years of age was not included in fat mass index measurements.

^d^ Beta estimate for cycle was .379 (P= .10) in this model (n = 1324). Data cycles of 2011-2018 were not included in this analysis because ≥60 years of age was not included in fat mass index measurements.

**Supplemental Table 15. Adjusted^a^ model-based means along with 95% confidence interval (CIs) for waist circumference among cancer survivors with non-missing fat mass index**

| **Data Cycle** | **Among ages 20–59 years^b^** | **Among ages ≥60 years^c^** | **Across All Ages^d^** |
| --- | --- | --- | --- |
| 1999-2000 | 93.9 (90.2-97.7) | 96.5 (93.0-100.1) | 96.8 (93.6-100.1) |
| 2001-2002 | 102.5 (98.9-106.2) | 100.5 (98.2-102.7) | 102.8 (100.7-104.8) |
| 2003-2004 | 102.1 (98.6-105.6) | 99.6 (97.3-102.0) | 101.8 (99.4-104.3) |
| 2005-2006 | 99.5 (96.1-102.9) | 99.8 (95.4-104.3) | 101.4 (98.8-104.1) |
| 2011-2012 | 101.3 (96.6-106.1) | NA | NA |
| 2013-2014 | 103.6 (99.3-107.9) | NA | NA |
| 2015-2016 | 103.3 (99.6-107.1) | NA | NA |
| 2017-2018 | 100.8 (95.8-105.8) | NA | NA |

NA, not available per NHANES survey age limitation for DEXA scans

^a^ The model included NHANES cycles, sex (male, female), age (continuous), and race/ethnicity category (non-Hispanic African American/Black, non-Hispanic White, Hispanic, and non-Hispanic other).

^b^ Beta estimate for cycle was .409 (P= .09) in this model (n = 801).

^c^ Beta estimate for cycle was 1.13 (P= .08) in this model (n = 891). Data cycles of 2011-2018 were not included in this analysis because ≥60 years of age was not included in fat mass index measurements.

^d^ Beta estimate for cycle **was 1.32 (P= .02)** in this model (n = 1238). Data cycles of 2011-2018 were not included in this analysis because ≥60 years of age was not included in fat mass index measurements.
